# Marital dissolution and cognition: The mediating effect of β-amyloid neuropathology

**DOI:** 10.1101/2024.05.15.24307413

**Authors:** Avinash Chandra, Rifah Anjum, Sheena Waters, Petroula Proitsi, Laura J Smith, Charles R Marshall, the Alzheimer’s Disease Neuroimaging Initiative

## Abstract

**Background:** Widowhood and divorce are extremely stressful life events and have been associated with high risk of dementia and cognitive impairment. However, the neurobiological mechanisms underlying how this risk is conferred requires further investigation. Alzheimer’s disease (AD) pathology, such as β-amyloid (Aβ), may explain influences of chronic stress, such as those seen in disruptive marital transitions, on declines in cognition. Therefore, we examined whether Aβ mediates associations between marital dissolution (through widowhood or divorce) and executive functioning (EF) and episodic memory (EM) performance in cognitively normal (CN) individuals.

**Methods:** Data from 543 CN participants from the Alzheimer’s Disease Neuroimaging Initiative (ADNI) were analysed. Outcomes included marital status, Aβ PET tracer uptake, and composite EF and EM scores. Primary analyses assessed relationships between marital dissolution and Aβ pathology, and marital dissolution and cognitive performance, and explored whether Aβ mediated associations between the latter.

**Results:** Marriage dissolution was associated with increased Aβ burden (β= 0.56; 95% CI: 0.11 to 1.02; *P*= 0.015) and worse EM performance (β= –0.09; 95% CI: –0.15 to –0.03; *P*= 0.003). Level of Aβ neuropathology was also identified as a significant mediator for the relationship between marriage dissolution and EM (ACME= –0.007; *P*= 0.029).

**Conclusions:** Aβ pathology was identified as a potential neurobiological mediator for the impacts of chronic stress due to marital dissolution on poorer memory performance. This suggests that stressful life events, such as the dissolution of one’s marriage might exert a direct effect on AD proteinopathy, which may subsequently influence poor cognition.

## Introduction

Major stress may adversely influence brain health and heighten the risk of dementia (1–3). Dissolution of marriage, through either divorce or death of a spouse, is common and has been identified as among the most stressful events across the adult lifespan (4). These experiences may result in poorer mental health and declines in health-related quality of life (5, 6). Moreover, marital dissolution is associated with an increased risk of cognitive impairment and dementia (7–14). The reasons for this are likely multifactorial and include limited access to shared economic and sociopsychological resources, encompassing social engagement and support, which promote better cognitive health after marital transitions (7). Adverse neurobiological impacts may also stem indirectly from the stress associated with marital disruption. For example, persistent stress may promote unhealthy behaviours and maladaptive coping such as excessive smoking and drinking which can lead to central nervous system (CNS) damage (7, 15, 16).

Stressful life events, such as marital dissolution, may have more direct effects on elements comprising the neuropathological cascade of Alzheimer’s disease (AD) (17), which is the most common cause of dementia (18). This includes toxic effects of chronic stress on hippocampal neurons (8, 19), neuroinflammation (20), focal neurodegeneration (20), and pathogenic protein accumulation, including β-amyloid (Aβ) aggregation (21). Aβ is thought to be a driving force facilitating this cascade (17). Whilst no direct relationship has yet been established between marital dissolution and Aβ pathology (22–25), previous research has found an interaction between amyloid and widowhood on cognitive decline. Widowed individuals that exhibited high levels of brain Aβ had steeper trajectories of cognitive decline (24). However, no study has yet examined mediating influences of cerebral Aβ levels on the relationship between both divorce and widowhood and domain-specific cognitive performance. Thus, the primary aim of the current study was to evaluate whether Aβ pathology measured through *in vivo* neuroimaging mediates influences of marriage dissolution on executive functioning (EF) and episodic memory (EM) performance in healthy older adults.

Exploring these effects in a healthy population could shed further light on the pathogenic mechanisms that underlie the role of psychosocial stress in contributing to the earliest signs of AD. These individuals are at a stage, well before the onset of any clinically meaningful symptomatology, when benefits of interventions and prevention strategies (e.g. social support, counselling, and access to community resources) may be at their most efficacious.

## Methods and materials

### Participants and data source

Data used in this study was obtained from the Alzheimer’s Disease Neuroimaging Initiative (ADNI; available at adni.loni.usc.edu). ADNI is a multi-centre and longitudinal study that prospectively recruits participants that are either cognitively normal (CN) or have diagnoses of mild cognitive impairment (MCI) or Alzheimer’s disease (AD). The primary aim of ADNI is the early identification of features associated with AD, in addition to assessing its progression, though the integration of neuroimaging measures and other biomarkers with cognitive and clinical information. The current investigation utilised cross-sectional data from 543 CN subjects from ADNI (Figure 1), who were either married or had their marriage dissolved (through divorce or death of spouse). Data was also available on age and cognitive performance at Aβ PET scan visit, sex, education level, and *Apolipoprotein E* (*APOE*) ε4 genotype.

**Figure 1.**
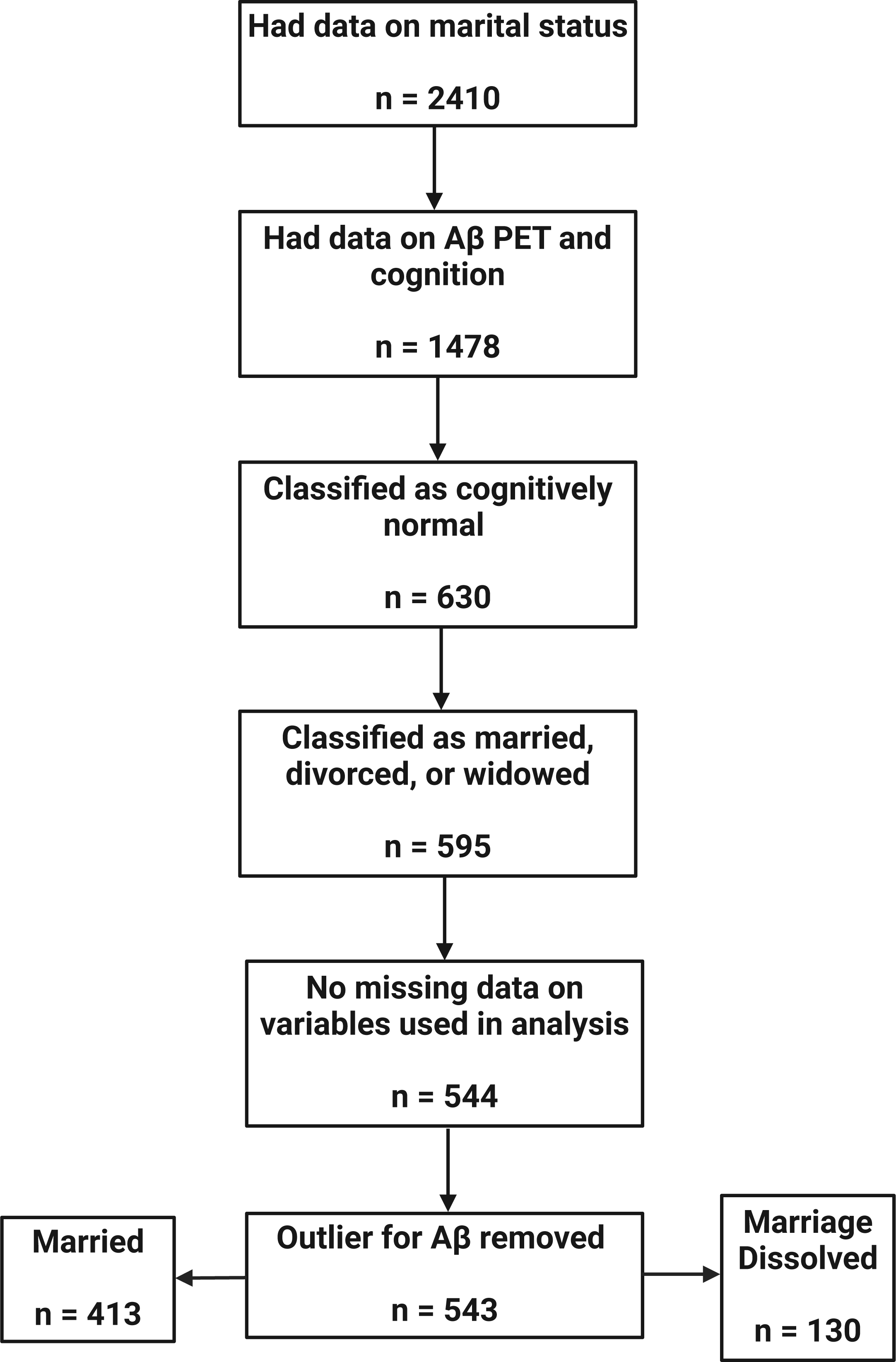
Sample size flowchart for ADNI subjects used in the current analysis. Created with BioRender.com.

### Measures

#### Marital status

Study participants reported their marital status at the time of the ADNI baseline visit. There were five response categories including: married, divorced, never married, unknown, and widowed. For current analyses, participants were categorised into one of two marital groups. The first were those who reported being married at baseline (control group), whilst the second were those who reported either being either widowed or divorced (‘marriage dissolved’ group).

#### Aβ PET imaging

Amyloid PET neuroimaging acquisition and preprocessing was undertaken within the ADNI study using either the radiotracers [^18^F]florbetapir (Amyvid) or [^18^F]florbetaben (Neuraceq). For [^18^F]florbetapir: participants were injected with a 370 ± 37 MBq bolus, and 3D amyloid PET scans were acquired 50–70 min post-injection, which lasted approximately 20 minutes and consisted of four X five-minute frames. For [^18^F]florbetaben: participants were injected with a 300 ± 30 MBq bolus, and 3D amyloid PET scans were acquired 90–110 min post-injection, which lasted approximately 20 minutes and consisted of four X five-minute frames. Pre-processing steps included magnetic resonance imaging (MRI) segmentation and parcellation using FreeSurfer 7.1.1, co-registration between respective MRI and PET images, intensity normalisation, and the calculation of standardised uptake value ratios (SUVR) including cortical summary SUVR values for both florbetapir and florbetaben. Further information on amyloid PET processing methods can be found on the ADNI website and previous work (26). Only scans that fully passed quality control and had no evidence of technical errors were selected for the current investigation.

Data from [^18^F]florbetapir and [^18^F]florbetaben was also utilised to calculate standardised Centiloid transformations by ADNI investigators, which allows for the direct comparison and harmonisation of Aβ levels derived from both radiotracers (27, 28). Briefly, direct normalised cortical SUVR to Centiloid unit conversion was conducted by adapting the level-2 analysis developed by Klunk and colleagues (29). Techniques utilised in this conversion included linear regressions, scaling, and transformation equations.

#### Cognition

Data on cognitive performance for the initial Aβ PET visit was quantified using a battery of standardised neuropsychological assessments. Cognitive tests utilised in the generation of the EF score included (1) Category Fluency – animals, (2) Category Fluency – vegetables, (2) Trail Making Test A, (3) Trail Making Test B, (4) Digit span backwards, (5) Wechsler Adult Intelligence Scale-Revised (WAIS-R) Digit Symbol Substitution, items from the (6) Clock Drawing test. Tests used for EM score generation included (1) Rey Auditory Verbal Learning Test (RAVLT), (2) Alzheimer’s Disease Assessment Scale–Cognitive Subscale (ADAS-Cog), (3) Wechsler Memory Scale-Revised (WMS-R) Logical Memory, and (4) recall tests from the Mini Mental Status Exam (MMSE).

Composite EF and EM scores were derived utilising item response theory methodology and had a mean of 0 and a standard deviation of 1. A more in-depth explanation for the processes and methods used to generate these composite scores can be found elsewhere (30, 31). These composite scores were chosen as they measure two neuropsychological domains that are typically impaired very early during mild AD, relative to other areas of cognition (e.g. language, sustained attention) (32).

#### Statistical analyses

The Shapiro-Wilk test and corresponding histograms were used to identify primary outcome variables with non-normal distributions. All three (Aβ Centiloid value, EF score, and EM score) met this criteria (Table S1, Figures S1-S3) and, due to the inclusion of negative values, were cube-root transformed to correct for this (33–35). Prior to transformation, outliers were detected using the ‘qqplot’ function in R (36) with one outlier being identified for Aβ Centiloid score and removed from analyses (Figure S4). Using absolute Z value computation (37), this case was additionally categorised as an extreme outlier with a Z value>5. In Aβ PET data using Centiloid values, quantification methods have been previously shown to influence the presence of outliers (38), which have been shown to impact sensitivity in neuroimaging modalities more broadly (39). Differences in demographic and baseline characteristics of the study sample split by marital status were conducted utilising independent samples t-tests for continuous variables and Chi-square tests for categorical variables.

Multiple linear regression was first run to assess the relationship between (1) marriage dissolution and Aβ levels and (2) marriage dissolution and cognition. Covariates for regressions with Aβ as the outcome included age, sex, education level, PET tracer, and *APOE* ε4 status, whilst those for regressions with cognition (EF or EM score) as the outcome included age, sex, and education level. For results that demonstrated significant associations, interactions between age, sex, education and/or *APOE* ε4 and marital dissolution were individually evaluated in separate secondary models (depending on the covariates utilised in initial models). This was done to examine whether associations between exposures and outcomes were different based on population characteristics. As part of this analysis, interactions were also tested between Aβ levels and marital dissolution to test for moderating or synergistic effects on cognition. Regression models with significant associations were selected for two-stage mediation analysis to determine whether Aβ pathology could mediate potential influences of marriage dissolution on cognition.

The Baron and Kenney (1986) criteria (40) were initially utilised to determine whether sufficient evidence was present for a mediation effect. Specifically, requirements were: (1) X (marriage dissolution) is correlated with Y (cognition), (2) X (marriage dissolution) is correlated with M (Aβ levels), (3) M (Aβ levels) is correlated with Y (cognition) - while controlling for X, and (4) the correlation between X (marriage dissolution) and Y (cognition) is reduced when including M (Aβ levels) in the model (partial mediation) or disappears (total mediation). The second stage of mediation analysis examined causal mediation effects using the ‘mediation’ R package (41). Specifically, we computed the average direct effect (ADE) and average causal mediation effect (ACME). These metrics respectively aim to quantify both direct and indirect mediated effects of Aβ levels on cognition. Non-parametric bootstrapping consisting of 5000 simulations was used to generate bias-corrected confidence intervals for ADE and ACME. Mediation analyses consisted of multiple regressions used in earlier analytic steps. Sensitivity analyses included re-testing for mediation and moderation effects (1) within stratified groups for covariates that demonstrated a significant interaction in secondary models and (2) when including both age and age squared in original regression models (fully adjusted model) to determine if results differ when accounting for non-linear effects of age (42). *P*<0.05 was set as the threshold for statistical significance across all models. All statistical analyses were performed using R software version 4.2.2 (R Project for Statistical Computing). This research utilised Queen Mary’s Apocrita HPC facility, supported by QMUL Research-IT. http://doi.org/10.5281/zenodo.438045, in addition to Open OnDemand (43).

## Results

### Demographic results

Samples characteristics can be found in Table 1. Groupwise comparison indicated that there were more women in the marriage dissolved category relative to men. Moreover, participants in this group also had higher levels of Aβ pathology relative to those that were married. Otherwise, no significant group differences were observed.

**Table 1.** Sample characteristics. This table presents demographic information and information on outcomes stratified by marital status.

| Variable | Married (n =413) | Marriage Dissolved (n =130) | P Value |
| --- | --- | --- | --- |
| Age at Aβ PET scan (years), M (SD) | 72.5 (6.6) | 73.8 (7.7) | 0.091 |
| Education (years), M (SD) | 16.7 (2.4) | 16.2 (2.6) | 0.068 |
| Sex, no.(% male) | 199 (48.2) | 33 (25.4) | <0.001*** |
| Race, no.(% white) | 376 (91) | 113 (86.9) | 0.230 |
| Time between baseline visit and Aβ PET Scan (months), M (SD) | 8.4 (23) | 4.9 (15.7) | 0.052 |
| APOE ε4 status, no.(%carrier)<sup>a</sup> | 131 (31.7) | 34 (26.2) | 0.274 |
| Aβ Centiloid level, M (SD) | 16 (31.7) | 24.2 (37.2) | 0.023* |
| Aβ PET tracer, no.(% [<sup>18</sup>F]florbetapir) | 310 (75.1) | 96 (73.8) | 0.871 |
| Executive functioning composite score, M (SD) | 0.8 (0.7) | 0.6 (0.7) | 0.066 |
| Episodic memory composite score, M (SD) | 1.1 (0.6) | 1.0 (0.6) | 0.129 |
| Marriage dissolution, no.(%widowed)<sup>b</sup> | - | 62 (47.7) | - |

<sup>a</sup>In the married group 121 heterozygous and 10 homozygous carriers; in the marriage dissolved group 32 heterozygous carriers and 2 homozygous carriers.
<sup>b</sup>Only applicable to marriage dissolution group. Pairwise comparison not possible. Values are relative to the divorced group.
\*Significant at $P<0.05$
\*\*\*Significant at $P<0.001$
Abbreviations: M, mean; SD, standard deviation; no., number

### Relationships between marriage dissolution, Aβ pathology, and cognition

Marriage dissolution was associated with increased levels of Aβ neuropathology quantified by standardised Centiloid values (β= 0.56; 95% CI: 0.11 to 1.02; *P*= 0.015; Figure 2). An association was also observed between marriage dissolution and lower EM scores (β= –0.09; 95% CI: –0.15 to –0.03; *P*= 0.003; Figure 3) but not between marriage dissolution and EF scores (β= –0.05; 95% CI: –0.16 to 0.05; *P*= 0.340; Figure 3). Through secondary models, an interaction effect between marriage dissolution and sex on higher EM score was observed (β= 0.20; 95% CI: 0.07 to 0.33; *P*= 0.003; Figure 4). This indicated that the effect of marital dissolution on worse EM is greater in men than in women. No other significant interaction effects were found, including between Aβ and marital dissolution (Table S2).

**Figure 2.**
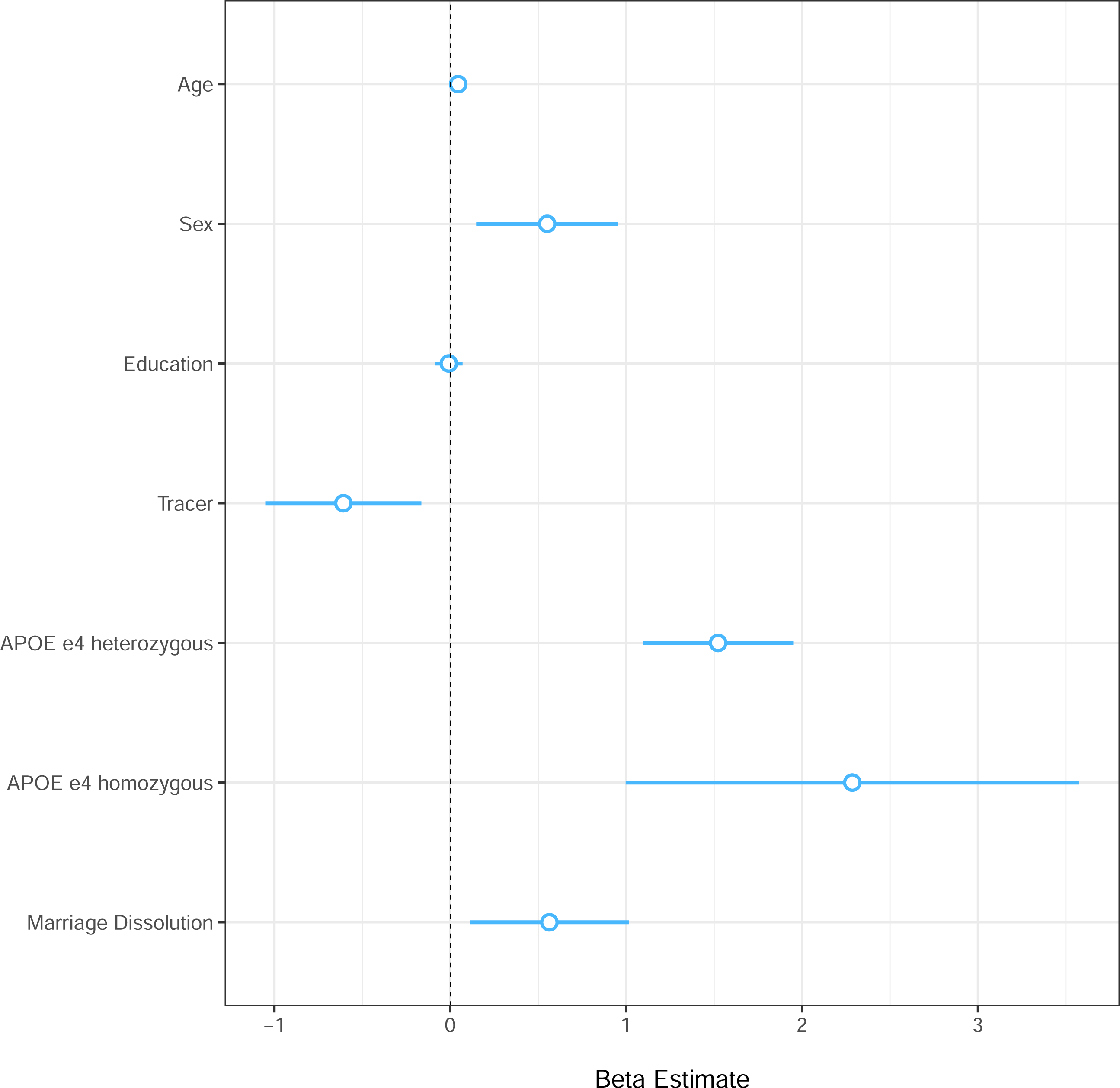
Associations between Aβ pathology and marriage dissolution. The forest plot displays the magnitude of regression coefficient estimates (β) for the model examining associations between marriage dissolution and Aβ neuropathology. Estimates on the x-axis represent β coefficients and error bars represent the 95% confidence intervals.

**Figure 3.**
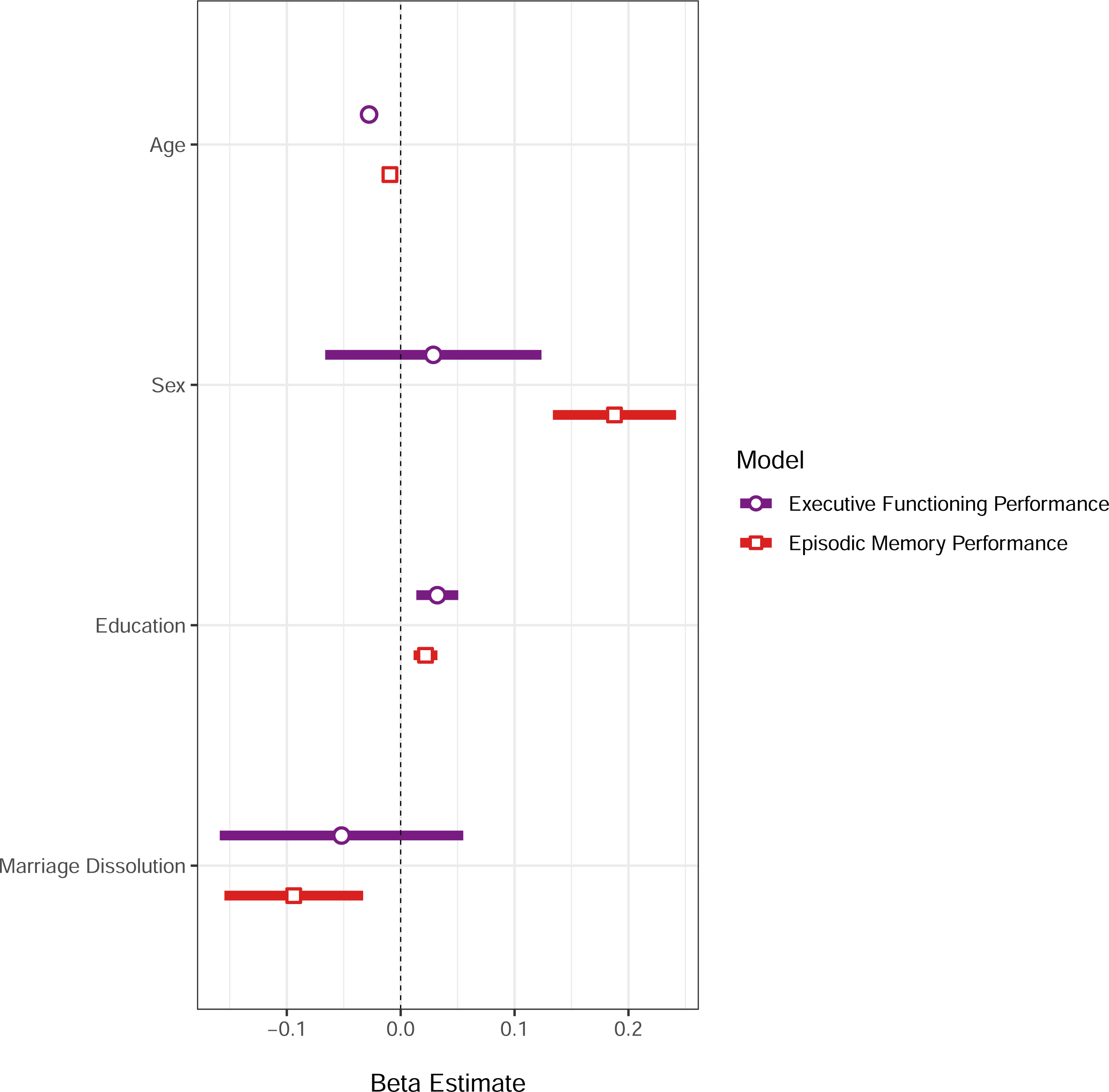
Associations between executive functioning and episodic memory performance and marriage dissolution. The forest plot displays the magnitude of regression coefficient estimates (β) for the 2 models examining associations between marriage dissolution and cognition. Estimates on the x-axis represent β coefficients and error bars represent the 95% confidence intervals.

**Figure 4.**
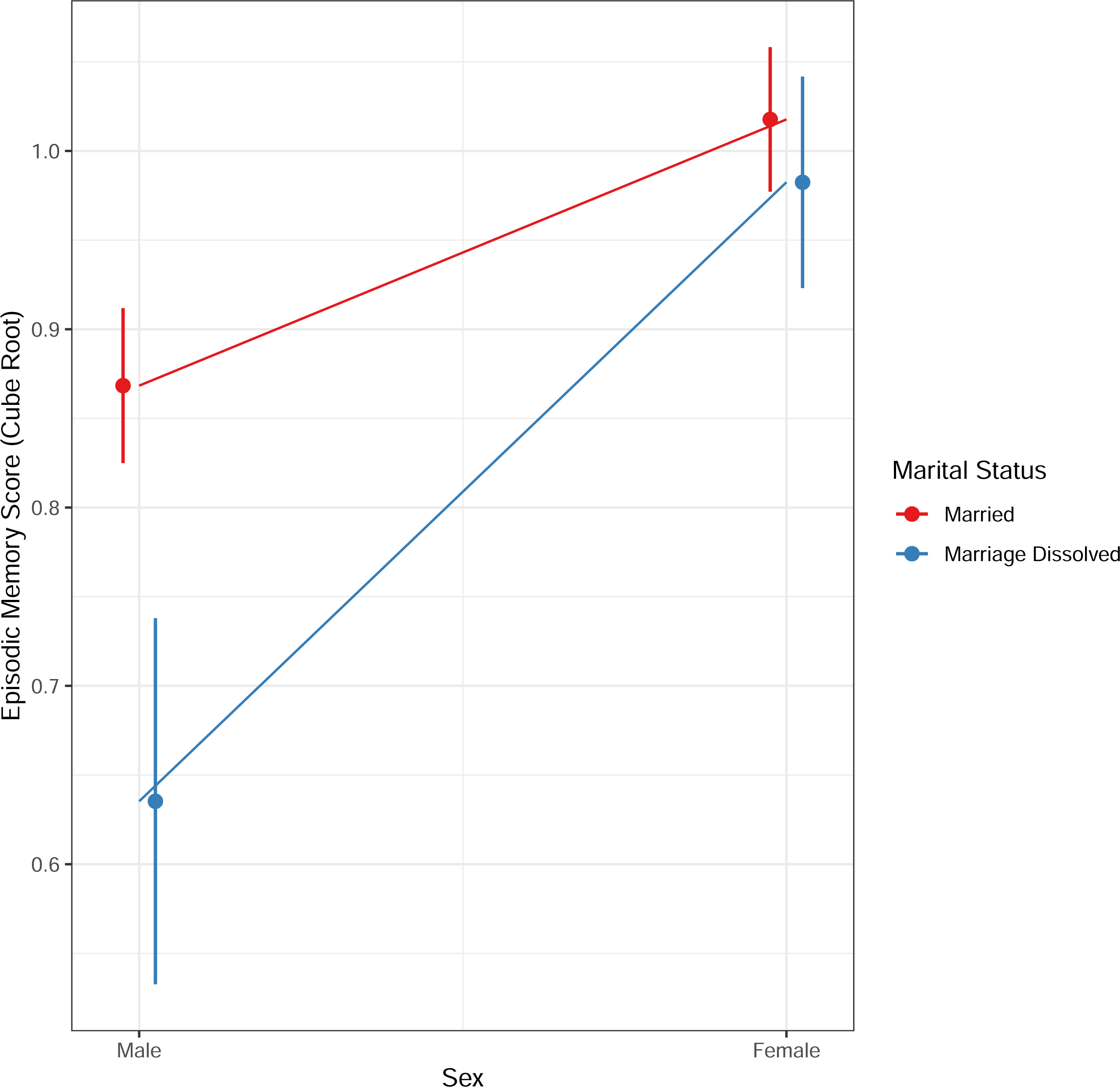
Interaction effect between marital dissolution and sex. The graphical plot indicates the magnitude and direction of effect of the significant interaction term (marital status*sex) from the model. Error bars represent the 95% confidence intervals.

### Mediation analyses

#### Baron and Kenney criteria for partial mediation

Given significant associations found between marriage dissolution, Aβ pathology and EM scores, mediating influences of Aβ on the relationship between marriage dissolution and memory were evaluated. As indicated previously, criteria 1 and 2 were met. Specifically, significant associations were observed between (1) marriage dissolution and memory performance and (2) marriage dissolution and Aβ pathology. Aβ levels were negatively associated (when including marital dissolution in the model) with memory scores (β= –0.01; 95% CI: –0.02 to –0.002; *P*= 0.021). After including Aβ pathology in the regression equation, the relationship between marriage dissolution and EM scores becomes weaker in terms of effect size and significance value (Table 2). Thus, criteria 3 and 4 of the Baron and Kenney criteria were met and sufficient evidence existed for a partial mediating effect of Aβ on the negative influence of marriage dissolution on memory performance.

**Table 2.** Mediation results using Baron and Kenney criteria. This table presents partial mediation results with and without Aβ as a covariate.

| Regression Model | $\beta^a$ | 95% Confidence Interval<sup>a</sup> | t-value | P Value |
| --- | --- | --- | --- | --- |
| (1) EM score ~ Marriage Dissolution | -0.094 | -0.155 to -0.033 | -3.03 | 0.003** |
| (2) EM score ~ Marriage Dissolution + Aβ | -0.088 | -0.149 to -0.027 | -2.83 | 0.005** |

<sup>a</sup>Rounded to 3 decimal places to demonstrate differences in effect size
\*\*Significant at $P<0.01$
Abbreviations: EM, episodic memory

#### Causal mediation effects

Results from non-parametric bootstrapping indicated that both direct and Aβ-mediated effects for the association between marriage dissolution and memory performance were present. Estimates for the average direct effect (*P*= 0.018) indicated a direct effect of marriage dissolution on memory scores when accounting for the mediation effect of Aβ, whilst those for the average causal mediation effect indicated a mediation or indirect (*P*= 0.029) effect of Aβ for this relationship (Table 3; Figure 5).

**Figure 5.**
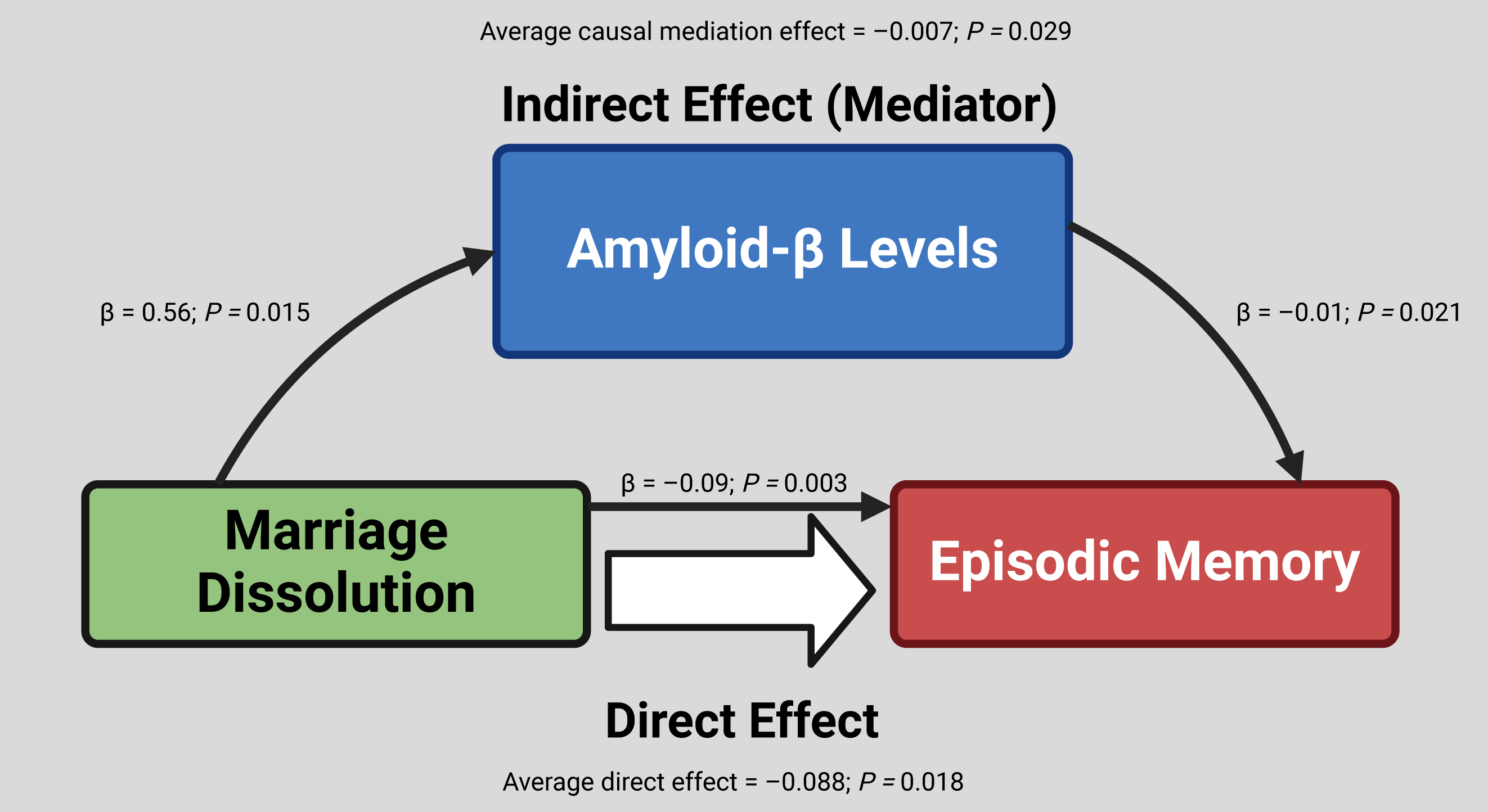
Diagram of mediation effects. The graphic indicates significant results from mediation analyses using the Barron and Kenney criteria and from causal mediation using non-parametric bootstrapping. Estimates and P values and corresponding lines are representative of individual regression models. Created with BioRender.com.

**Table 3.** Mediation results using bootstrapping. This table presents results from mediation results using nonparametric bootstrap confidence intervals with the percentile method.

| Outcome | Estimate<sup>a</sup> | 95% Confidence Interval<sup>a</sup> | P Value |
| --- | --- | --- | --- |
| Average causal mediation effect | −0.007 | −0.017 to −0.0004 | 0.029<sup>*</sup> |
| Average direct effect | −0.088 | −0.167 to −0.016 | 0.018<sup>*</sup> |
| Total effect | −0.095 | −0.173 to −0.024 | 0.009<sup>**</sup> |
| Proportion Mediated | 0.075 | 0.002 to 0.356 | 0.038<sup>*</sup> |

<sup>a</sup>Rounded to 3 decimal places to highlight precise effect size
<sup>\*</sup>Significant at $P<0.05$
<sup>\*\*</sup>Significant at $P<0.01$
Abbreviations: EM, episodic memory

#### Sensitivity analyses

When stratifying the sample by sex, no significant interaction effects were found between Aβ and marital dissolution for males (β= 0.03; 95% CI: –0.03 to 0.08; *P*= 0.321) or females (β= –0.02; 95% CI: –0.05 to 0.01; *P*=0.154) on memory scores. Moreover, no significant mediation effects were observed when stratifying by sex. Specifically, for males, no significant association between marriage dissolution and Aβ pathology (Criteria 2; β= 0.41; 95% CI: –0.47 to 1.29; *P*= 0.358) or between Aβ pathology and memory performance when controlling for marital dissolution (Criteria 3; β= –0.02; 95% CI: –0.04 to 0.003; *P*= 0.092) were found. For females, no significant associations were found between marriage dissolution and memory performance (Criteria 1; β= –0.04; 95% CI: –0.10 to 0.02; *P*= 0.163) or between Aβ pathology and memory performance when controlling for marital dissolution (Criteria 3; β= –0.01; 95% CI: –0.02 to 0.003; *P*= 0.143).

In fully adjusted models (accounting for age squared), Baron and Kenney criteria for partial mediation was still met. Specifically, significant associations were observed between marriage dissolution and memory performance (Criteria 1; β= –0.09; 95% CI: –0.15 to –0.03; P= 0.003), marriage dissolution and Aβ pathology (Criteria 2; β= 0.59; 95% CI: 0.14 to 1.05 *P*= 0.011), and Aβ levels and memory scores when including marital dissolution as a covariate (Criteria 3; β= – 0.01; 95% CI: –0.02 to –0.002; *P*= 0.021). When including Aβ pathology in the regression equation, the relationship between marriage dissolution and EM scores becomes weaker in terms of effect size and significance value (Criteria 4; Table 4). Results from non-parametric bootstrapping confirmed that both direct and Aβ-mediated effects for the association between marriage dissolution and memory performance were still present when including age squared into models (Table 5).

**Table 4.**
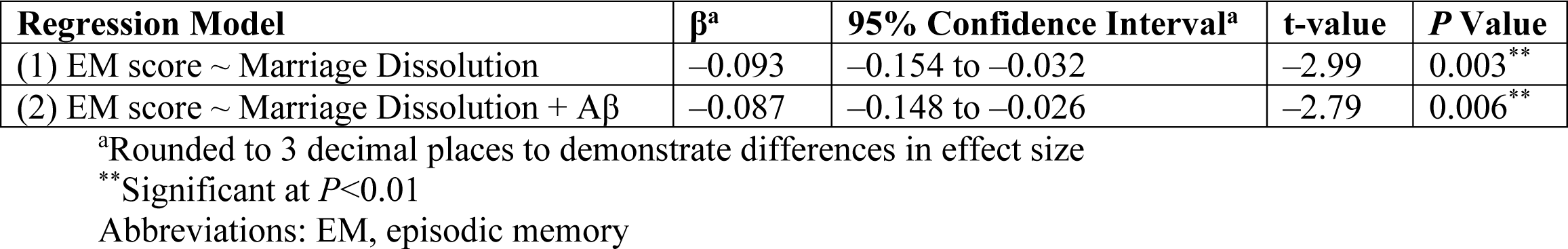
Mediation results using Baron and Kenney criteria including age squared. This table presents partial mediation results with and without Aβ as a covariate with age squared included in models.

| Regression Model | $\beta^a$ | 95% Confidence Interval<sup>a</sup> | t-value | P Value |
| --- | --- | --- | --- | --- |
| (1) EM score ~ Marriage Dissolution | −0.093 | −0.154 to −0.032 | −2.99 | 0.003<sup>**</sup> |
| (2) EM score ~ Marriage Dissolution + $A\beta$ | −0.087 | −0.148 to −0.026 | −2.79 | 0.006<sup>**</sup> |

<sup>a</sup>Rounded to 3 decimal places to demonstrate differences in effect size
<sup>\*\*</sup>Significant at $P<0.01$
Abbreviations: EM, episodic memory

**Table 5.** Mediation results using bootstrapping including age squared. This table presents results from mediation results using nonparametric bootstrap confidence intervals with the percentile method with age squared included in models.

| Outcome | Estimate<sup>a</sup> | 95% Confidence Interval<sup>a</sup> | P Value |
| --- | --- | --- | --- |
| Average causal mediation effect | −0.008 | −0.018 to −0.001 | 0.024<sup>*</sup> |
| Average direct effect | −0.087 | −0.165 to −0.015 | 0.019<sup>*</sup> |
| Total effect | −0.094 | −0.172 to −0.022 | 0.009<sup>**</sup> |
| Proportion Mediated | 0.080 | 0.003 to 0.373 | 0.034<sup>*</sup> |

<sup>a</sup>Rounded to 3 decimal places to highlight precise effect size
<sup>\*</sup>Significant at $P<0.05$
<sup>\*\*</sup>Significant at $P<0.01$
Abbreviations: EM, episodic memory

## Discussion

Here we have shown that marital dissolution is associated with poorer episodic memory function, and this association is mediated by Aβ burden. We evaluated relationships between marriage dissolution through either divorce or death of a spouse, Aβ levels quantified by Centiloid values, and cognitive performance on EF or EM, using a two-stage analysis to determine a mediation effect for Aβ burden. Mediation results were robust to potential non-linear influences of age and were not observed when stratifying marital groups by sex, potentially due to loss of power. Overall, these findings further validate the importance of divorce and widowhood as risk factors for the development of cognitive impairment, but go further than previous studies by demonstrating that this is partially mediated by influences of neuropathological change characteristic of AD, namely elevated CNS Aβ neuropathology.

Our findings complement previous research that identified *in vivo* Aβ pathology as a moderator of widowhood and cognitive decline. Prior work from Biddle and colleagues (2022) suggested that the experience of spousal death could interact or exert parallel influences with Aβ burden to exacerbate cognitive deterioration (24). However, this result was not observed in the current investigation using data from ADNI. Whilst we adapted a cross-sectional approach, the present study provides the first justification that Aβ burden may mediate the relationship between the stress affiliated with both loss of a spouse through death or divorce and poorer performance on EM, which is one of the earliest clinical markers prior to the onset of AD (44). High levels of uncontrollable chronic stress, potentially attributable to factors like low levels of life satisfaction and unhappiness experienced after marital dissolution (45), can have more immediate neurobiological effects (46) that may explain the current results.

Preclinical research suggests that stress-induced alterations in brain immune response can result in microglial dysfunction, which may in turn lead to an aggregation of CNS Aβ pathology through impairments in degradation and clearance (47). Furthermore, it was evidenced in non-human primates that continuous glucocorticoid administration, typically released during stress, facilitated increases in Aβ_42_ plasma levels and reductions in insulin-degrading enzyme mRNA (48). Several studies utilising AD mouse models have consistently shown that exposure to long-term stress exacerbates the deposition of Aβ plaques in addition to interstitial Aβ (49–52), with corticosterone possibly intermediating these effects (52). Interestingly, we found no significant effects with respect to EF performance. EF has been linked to brain areas belonging executive control network (ECN), whereas EM to those that comprise the default mode network (DMN) (53). Research has shown that in even in MCI, ECN connectivity is not disrupted (54). However, even in cognitively healthy individuals, disruptions in the DMN, linked to memory decline, may be attributable, at least in part, to early Aβ neuropathology (55). This may be due to neurotoxic effects of Aβ on metabolic processes within afflicted neurons (56). Within the current study, influences of chronic stress on Aβ may have in turn impacted memory performance through DMN alterations.

To our knowledge, this study provides the first evidence of a neurobiological mediator between marital dissolution (through either widowhood or divorce) and cognitive performance. This adds to other possible mediators of this relationship identified in the literature including mental and physical health (57), number of children (58), and social integration and financial resources (59). This study also highlights the importance of stressful marital transitions as a risk factor for cognitive decline and potentially AD. Whilst widowhood and divorce have not been explicitly identified as one of the major modifiable risk factors for dementia, their experience may increase the frequency and intensity of a constellation of unhealthy behaviours identified by the Lancet commission (60). These include, but are not limited to, excessive smoking and drinking as deleterious coping strategies for these stressful life events, increased symptoms of depression, social withdrawal and isolation after spousal loss, and increased risk of diabetes and cardiovascular illnesses (7, 61–63). Moreover, our findings related to Aβ pathology imply that dementia risk cannot be wholly ascribed to these other risk factors. Marital dissolution may warrant consideration as a life course dementia risk factor in its own right.

Statistics demonstrate an increasing rate of divorce (64), and global projections suggest increases in advanced ageing (65), likely indicating a higher future prevalence of widowhood. Thus, offering these individuals resources to cope with the stress of marital transitions may be justified as a public health priority to promote cognitive health and well-being. This may take the form of support groups to encourage social engagement and alleviate loneliness (66), dietary strategies to target co-morbidities (67), or interpersonal psychotherapy to manage bereavement and loss (68). Findings from the current work also suggest that this group may warrant consideration as a priority for access to emerging anti-amyloid disease modifying therapies for Alzheimer’s disease (69).

Secondary results indicated that among individuals who had their marriage dissolved, women had higher EM performance than men. This finding is consistent with studies which found that divorce was more strongly associated with a higher risk of dementia for men than for women in Sweden (70) and in the general U.S. population (7). Based on existing data (71, 72), future studies may be warranted to more comprehensively examine whether the loss of a spouse may hurt men’s health and well-being, including cognitive health, more than women’s. Additionally, this may shed light on whether men, more than woman, are likely to benefit from interventions designed to counteract the adverse psychological effects of losing a spouse. However, there was also a noticeable underrepresentation of men in the marital dissolution group, which raises the possibility of ascertainment bias (73) during data collection in ADNI that may have influenced this finding. We aimed to address this through sex-stratified analyses.

Despite its novelty, the current study has several limitations that need to be addressed. Firstly, we utilised a cross-sectional study design, so temporal influences of marital dissolution could not be assessed. Moreover, as the ADNI study excluded subjects with Major Depressive Disorder or significant depressive symptomatology (Geriatric Depression Scale (GDS) Score>6), we did not include any measures of depression. A lack of such psychological measures makes it difficult to directly test our stress hypothesis or loss of social support in explaining our results. Our sample was also limited in terms of race, with over 90% consisting of White participants. These results require validation in cohorts consisting of a range of ethnicities including, but not limited to, Black and South Asian individuals who may be at elevated risk of dementia and have a younger age of dementia onset (74) and individuals from different cultural backgrounds who may have varying familial support structures. Moreover, whilst we combined widowed and divorced individuals into one group due to the hypothesised shared stress, there may be key differences among these groups. These include older ages for widows/widowers, additional psychological burden in the widowhood group due to bereavement, and the unique financial impacts of divorce. Data on marital status was also only collected at one timepoint during study entry. Although the number of average months between baseline and the initial amyloid PET scan visits was low, it is possible that some participants who were initially married at baseline had lost their partners by the time of their scan. Further research could also explore additional factors such as finding a new partner after spousal loss, quality of the marriage prior to dissolution, or duration of time after widowhood or divorce.

## Data Availability

The ADNI dataset can be accessed by approved users on https://adni.loni.usc.edu

## Acknowledgements

Data collection and sharing for the Alzheimer’s Disease Neuroimaging Initiative (ADNI) is funded by the National Institute on Aging (National Institutes of Health Grant U19 AG024904). The grantee organization is the Northern California Institute for Research and Education. In the past, ADNI has also received funding from the National Institute of Biomedical Imaging and Bioengineering, the Canadian Institutes of Health Research, and private sector contributions through the Foundation for the National Institutes of Health (FNIH) including generous contributions from the following: AbbVie, Alzheimer’s Association; Alzheimer’s Drug Discovery Foundation; Araclon Biotech; BioClinica, Inc.; Biogen; Bristol-Myers Squibb Company; CereSpir, Inc.; Cogstate; Eisai Inc.; Elan Pharmaceuticals, Inc.; Eli Lilly and Company; EuroImmun; F. Hoffmann-La Roche Ltd and its affiliated company Genentech, Inc.; Fujirebio; GE Healthcare; IXICO Ltd.; Janssen Alzheimer Immunotherapy Research & Development, LLC.; Johnson & Johnson Pharmaceutical Research &Development LLC.; Lumosity; Lundbeck; Merck & Co., Inc.; Meso Scale Diagnostics, LLC.; NeuroRx Research; Neurotrack Technologies; Novartis Pharmaceuticals Corporation; Pfizer Inc.; Piramal Imaging; Servier; Takeda Pharmaceutical Company; and Transition Therapeutics.

