## Supplementary Material for "Marital dissolution and cognition: The mediating effect of β-amyloid neuropathology"

**Table S1.** Results from normality testing. This table presents results from Shapiro-Wilk tests of normality.

| **Variable** | **Shapiro-Wilk Value (W)** | ***P* Value** |
|---|---|---|
| Aβ Centiloid value | 0.82 | <2.2e-16*** |
| EF Score | 0.99 | 0.0008*** |
| EM Score | 0.99 | 0.005** |

***Significant at *P*<0.001
**Significant at *P*<0.01
Significant value indicates violation of normality assumption
Abbreviations: EF, executive functioning; EM, episodic memory.

**Figure S1.** Frequency distribution for amyloid data. This histogram indicates the distribution of Aβ Centiloid values used in combination with the Shapiro-Wilk tests to identify non-normal distributions.

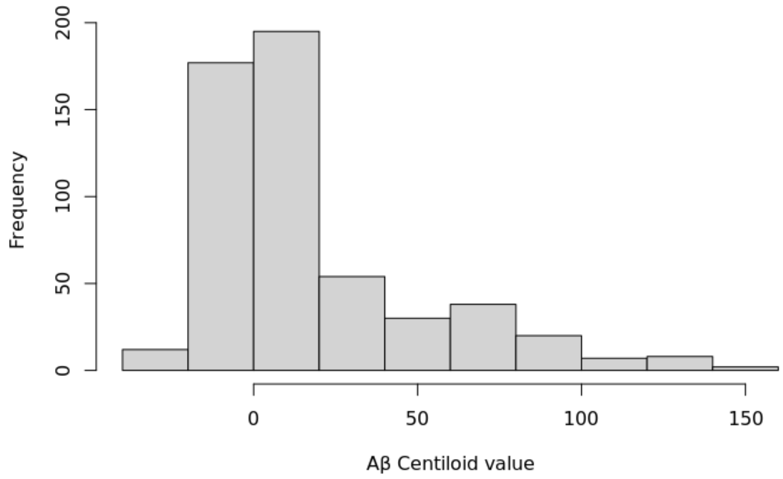

**Figure S2.** Frequency distribution for executive functioning performance data. This histogram indicates the distribution of executive functioning scores used in combination with the Shapiro-Wilk tests to evaluate normality of distributions.

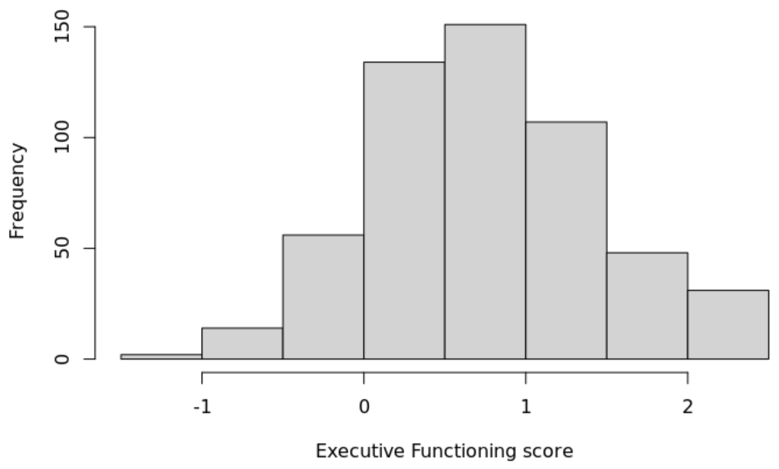


**Figure S3.** Frequency distribution for episodic memory performance data. This histogram indicates the distribution of episodic scores used in combination with the Shapiro-Wilk tests to evaluate normality of distributions.

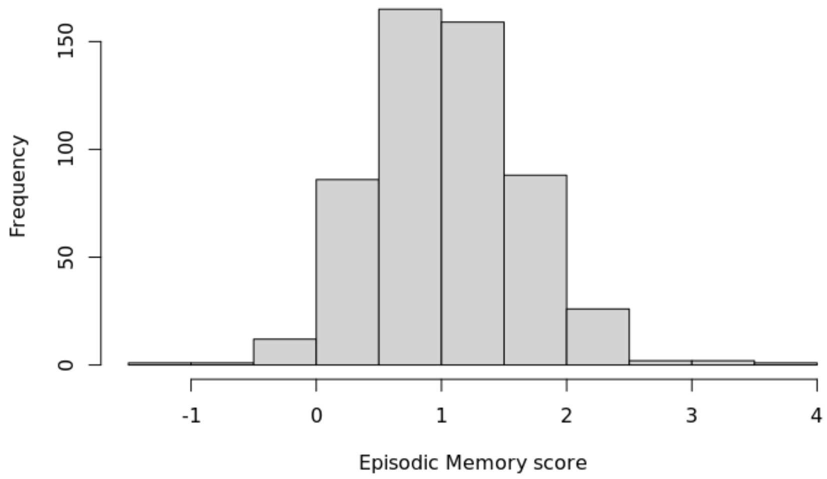

**Figure S4.** QQ plot for amyloid data. This QQ plot indicates the presence of an extreme outlier for Aβ Centiloid values, which was subsequently removed.

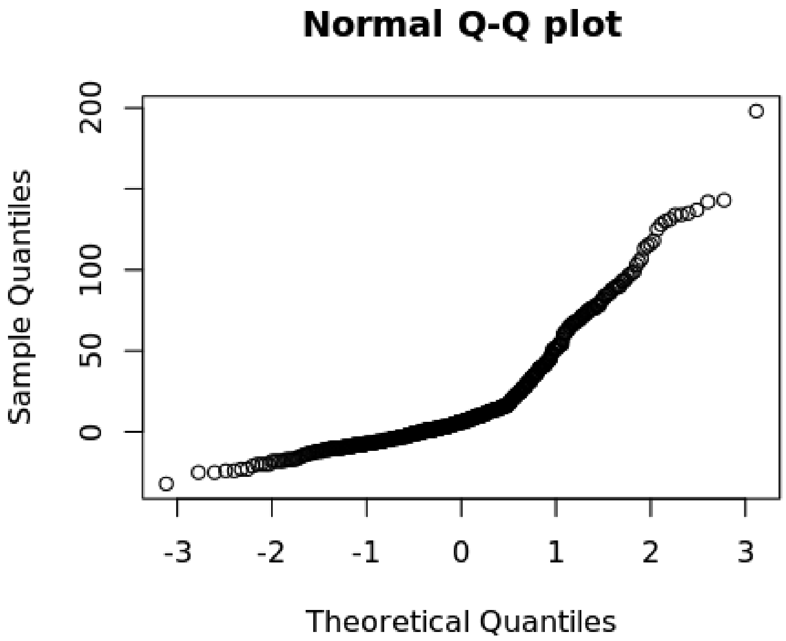


**Table S2.** Secondary models with interaction terms. This table presents results from analyses examining influence interaction terms that include marital dissolution on (1) Aβ and (2) episodic memory performance.

| **Regression Model** | **Interaction Term** | **β** | **95% Confidence Interval** | **t-value** | ***P* Value** |
|---|---|---|---|---|---|
| Model 1: Aβ | Marriage Dissolution * Age | –0.02 | –0.08 to 0.04 | –0.56 | 0.574 |
| Model 2: Aβ | Marriage Dissolution * Sex | 0.19 | –0.79 to 1.18 | 0.39 | 0.700 |
| Model 3: Aβ | Marriage Dissolution * Education | –0.04 | –0.22 to 0.13 | –0.48 | 0.628 |
| Model 4: Aβ | Marriage Dissolution * *APOE* ε4 (heterozygous) | 0.66 | –0.35 to 1.68 | 1.28 | 0.201 |
| Model 4: Aβ | Marriage Dissolution * *APOE* ε4 (homozygous) | 0.96 | –2.47 to 4.40 | 0.55 | 0.581 |
| Model 5: EM score | Marriage Dissolution * Age | –0.004 | –0.01 to 0.004 | –1.07 | 0.285 |
| Model 6: EM score | Marriage Dissolution * Sex | 0.20 | 0.07 to 0.33 | 2.96 | $0.003^{**}$ |

| | | | | | |
|---|---|---|---|---|---|
| Model 7: EM score | Marriage Dissolution * Education | 0.003 | –0.02 to 0.03 | 0.28 | 0.780 |
| Model 8: EM score | Marriage Dissolution * Aβ | 0.003 | –0.02 to 0.03 | 0.26 | 0.794 |

**Significant at $P<0.01$

Abbreviations: EM, episodic memory
